# National Trends in Mortality and Urgent Dialysis after Admission for Acute Hypertension in Japan from 2010 through 2019

**DOI:** 10.1101/2023.08.07.23293627

**Authors:** Hisazumi Matsuki, Taku Genma, Shintaro Mandai, Tamami Fujiki, Yutaro Mori, Fumiaki Ando, Takayasu Mori, Koichiro Susa, Soichiro Iimori, Shotaro Naito, Eisei Sohara, Tatemitsu Rai, Kiyohide Fushimi, Shinichi Uchida

**Affiliations:** Department of Nephrology, Graduate School of Medical and Dental Sciences, Tokyo Medical and Dental University, 1-5-45 Yushima, Bunkyo, Tokyo 113-8519, Japan; Department of Nephrology and Hypertension, Dokkyo Medical University, 880 Kitakobayashi, Mibu, Shimotsuga, Tochigi, 321-0293, Japan; Department of Health Policy and Informatics, Graduate School of Medical and Dental Sciences, Tokyo Medical and Dental University, 1-5-45 Yushima, Bunkyo, Tokyo 113-8519, Japan

**Keywords:** acute hypertension, dialysis, malignant hypertension, hypertensive emergency

## Abstract

**Background:** Despite increasing incidences of hypertension, recent trends in mortality and urgent dialysis following acute hypertension (AHT) remain undetermined.

**Methods:** This retrospective observational cohort study evaluated 50,316 hospitalized AHT patients from 2010 to 2019, using an administrative claims database in Japan. We examined trends in incidence, urgent dialysis, mortality and its risk factors using logistic regression models. Using ICD-10 codes, AHT was categorized into five spectrums: malignant hypertension (MHT) (*n* = 1,792), hypertensive emergency (*n* = 17,907), hypertensive urgency (*n* = 1,562), hypertensive encephalopathy (*n* = 6,593), and hypertensive heart failure (HHF) (*n* = 22,462).

**Results:** The median age of the patients was 76 years and 54.9% were female. The total AHT incidence was 70 cases per 100,000 admission year. The absolute death rate increased from 1.83% [95% confidence intervals (CI), 1.40–2.40] to 2.88% (95%CI, 2.42–3.41) [Cochran-Armitage trend test (CA), *P* < 0.0001]. Upward trends were observed in patients aged ≥80, with lean body mass index ≤18.4, and with HHF. Urgent dialysis rates increased from 1.52% (95%CI, 1.12–2.06) to 2.60% (2.17–3.1) (CA, *P* = 0.0071) in 48,235 patients, excluding maintenance dialysis patients. Older age, male, lean body mass, MHT, HHF, and underlying chronic kidney disease (CKD) correlated with higher mortality risk; greater hospital volume correlated with lower mortality risk; and MHT, HHF, diabetes mellitus, CKD, and scleroderma correlated with a higher risk of urgent dialysis.

**Conclusions:** Mortality and urgent dialysis rates following AHT have increased. Aging, complex comorbidities, and HHF-type AHT contributed to the rising trend of mortality.

## Non-standard Abbreviations and Acronyms

AKI Acute kidney injury

BMI Body mass index

CI Confidence intervals

CKD Chronic kidney disease

ESKD End-stage kidney disease

HHF Hypertensive heart failure

OR Odds ratios

SLE Systemic lupus erythematosus

## Introduction

Hypertension remains a substantial burden on the health of individuals worldwide. The number of adults with hypertension doubled from 1990 to 2019 globally, reaching nearly 1,300 million adults.^1^ Despite recent advances and diverse options for treating hypertension and more universal coverage of hypertensive patients worldwide, associated mortality and healthcare costs have increased as a result of acute or chronic systemic organ damage, including heart failure, stroke, and kidney insufficiency.^1–3^

Malignant hypertension (MHT) or hypertensive emergencies represent an accelerating rise in blood pressure with single or multiple organ failure. The related spectrums are known as acute hypertension (AHT), because of the heterogenicity of clinical presentation, severity, and blood pressure.^4, 5^ The incidence of AHT is less common, but the mortality rate is much higher compared with common hypertension; however, the epidemiology of community-dwelling patients with AHT is usually based on studies over a relatively short-term period in a small population.^6–8^ These studies have shown that the incidence of hospitalization or visits to emergency departments resulting from AHT have substantially increased, whereas trends in mortality have relied upon the limited studies and are devoid of recent updates over the past decade.^9^ In addition, the rate of urgent dialysis following hospitalization for AHT and the contribution to clinical outcome has not been determined.

Therefore, we examined the recent trends in incidence rates, mortality, and urgent dialysis of AHT from 2010 through 2019, before the COVID-19 pandemic, in a national population-based study using an administrative claims database in Japan. This study provides clarity to the increasing trends in mortality and urgent dialysis following admission for AHT as well as the contributing factors.

## Materials and Methods

### Source of data

We recruited study participants from the Diagnosis Procedure Combination inpatient database in Japan from 2010 to 2019. Details of this database have been described previously.^10–12^ Briefly, this national administrative claims database includes cases from 82 teaching hospitals and more than half of the hospitals in Japan. Nearly 7 million cases are included annually in the dataset. The database includes diagnoses and comorbidities at hospital admission and deaths as well as patient information, such as age, sex, body mass index (BMI), and patient care processes, including surgical procedures or used devices.

There were 72,334,105 hospital admissions from 2010 to 2019. Inclusion criteria in the analyses were age ≥18 years, hospitalization primarily for acute hypertension, and patients without pregnancy. Participants who died <24 h after admission were excluded.

Individuals with end-stage kidney disease (ESKD) undergoing maintenance hemodialysis and peritoneal dialysis were identified based on coding of patient care procedures as follows: chronic maintenance hemodialysis with <4 hours per session, ≥4 hours and <5 hours per session, ≥5 hours per session, chronic maintenance hemodiafiltration, or continuous peritoneal dialysis.^12, 13^ Urgent dialysis was recognized if patients received their initial dialysis during hospitalization for AHT.^12^

The ethics committee of the Tokyo Medical and Dental University approved this study. The need for informed consent was waived because of data anonymity. This study was performed in accordance with the ethical principles of the Declaration of Helsinki.

### Patient characteristics and outcome

AHT was defined and classified into five clinical spectrums based on the International Classification of Disease and Related Health Problems, 10th Revision (ICD-10)^14^ as follows: malignant hypertension (MHT) (I10), hypertensive emergency (I10), hypertensive urgency (I10), hypertensive encephalopathy (I674), and hypertensive heart failure (HHF) (I110).

Patient-level variables included age, sex, BMI, updated Charlson comorbidity index (CCI),^15^ and year of admission for the analyses. In particular, we identified comorbidities related to secondary hypertension, including diabetes mellitus, chronic kidney disease (CKD), maintenance dialysis-dependent ESKD, sleep apnea syndrome, pheochromocytoma, primary aldosteronism, renal artery stenosis, Takayasu arteritis, scleroderma, and systemic lupus erythematosus (SLE), as shown in Table S1. Hospital-level variables included hospital volume, which was defined as the mean number of daily hospitalized patients.^13^

The primary outcome was the occurrence of in-hospital death from any cause. The secondary outcome was the need for urgent dialysis during hospitalization for AHT. Patients were followed up until discharge, transfer, or death.

### Data analyses

Patient demographics and characteristics are presented as numbers and percentages or medians with interquartile range (IQR). The incidence of hospitalization resulting from AHT was calculated using the absolute number of admission cases for AHT divided by the total number of admission cases for any cause that were registered in the database in each year. The Cochran-Armitage test was used to evaluate the trend of absolute death rates and urgent dialysis rates. Multivariable logistic regression models were used to estimate the risk factors for all-cause mortality in whole groups and those for urgent dialysis in a subpopulation, excluding patients receiving maintenance dialysis upon admission. Potential confounding variables, including age, sex, BMI (≤18.4, 18.5–24.9, ≥25.0 kg/m^2^), hospital volume (tertiles), year of admission, AHT spectrum, and CCI (Model 1) or comorbidities related to secondary hypertension (Model 2) were adjusted. To clarify the impact of urgent dialysis on mortality, logistic regression models were used. The results are presented as odds ratios (OR) with 95% confidence intervals (CIs). Statistical analyses were performed using Stata version 15.0 software (Stata Corp., College Station, TX, USA). *P* values of <0.05 were considered statistically significant.

## Results

### Patient characteristics

This study enrolled 50,316 adults who required hospitalization for AHT. The participants were divided into five clinical categories including MHT (*N* = 1,792), hypertensive emergency (*N* = 17,907), hypertensive urgency (*N* = 1,562), hypertensive encephalopathy (*N* = 6,593), and HHF (*N* = 22,462). Table 1 lists the demographics and characteristics of the patients. The patients in the MHT group were markedly younger, the proportion of male patients was higher, and they were more obese compared with the other four groups. The proportion of lean body mass, diabetes mellitus, and greater general comorbidities were higher in the HHF group. The MHT patients exhibited a higher proportion of underlying secondary hypertension, including autoimmune and primary glomerular diseases. CKD was most common in MHT, followed by HHF, whereas that was the least common in hypertensive encephalopathy. Hypertensive retinopathy was more common in MHT and hypertensive urgency, and less common in the other types.

**Table 1.**
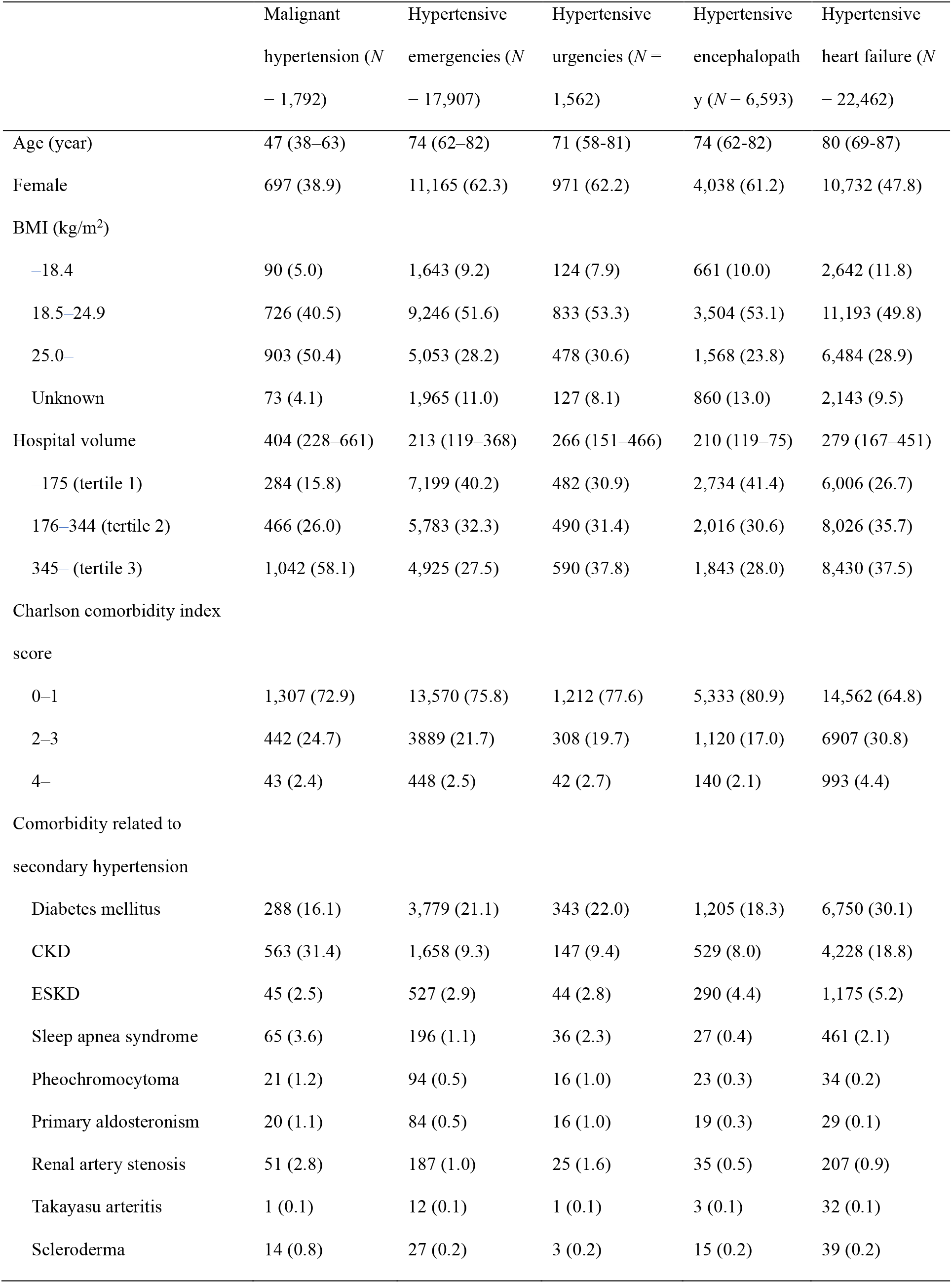
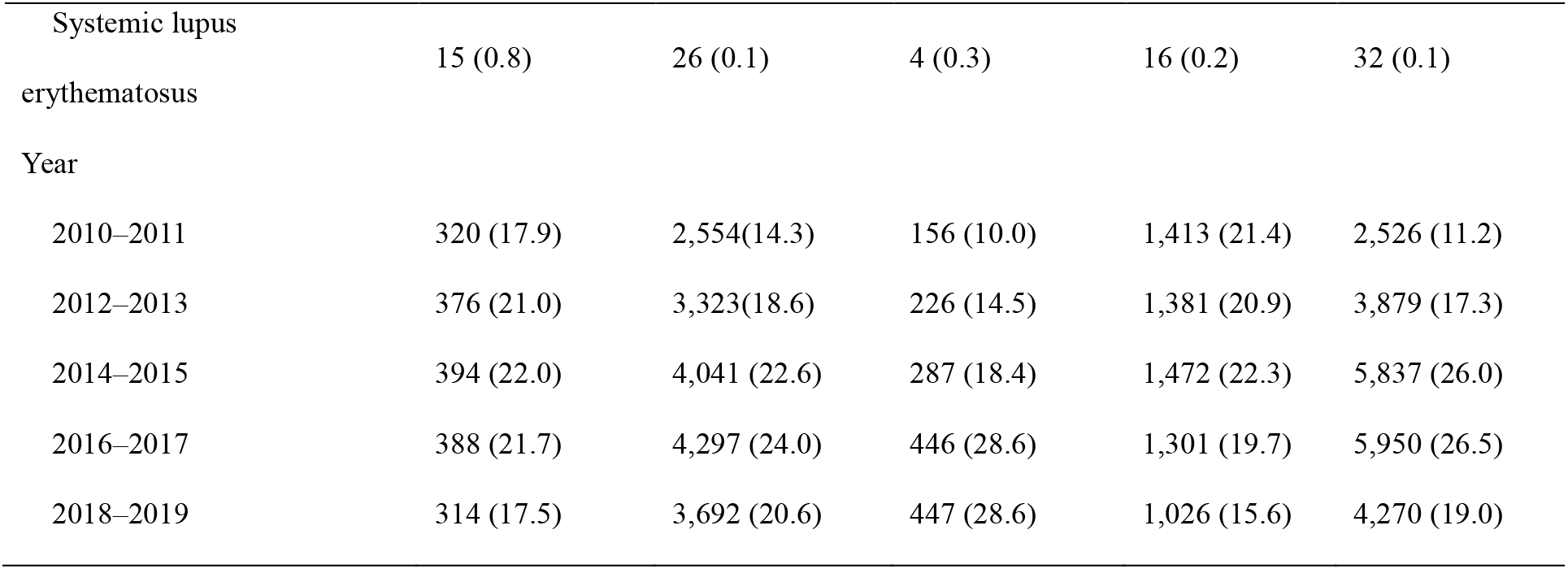
Characteristics of adults admitted with a primary diagnosis of acute hypertension.

|  | Malignant hypertension (*N* = 1,792) | Hypertensive emergencies (*N* = 17,907) | Hypertensive urgencies (*N* = 1,562) | Hypertensive encephalopathy (*N* = 6,593) | Hypertensive heart failure (*N* = 22,462) |
| --- | --- | --- | --- | --- | --- |
| Age (year) | 47 (38–63) | 74 (62–82) | 71 (58–81) | 74 (62–82) | 80 (69–87) |
| Female | 697 (38.9) | 11,165 (62.3) | 971 (62.2) | 4,038 (61.2) | 10,732 (47.8) |
| BMI (kg/m<sup>2</sup>) |  |  |  |  |  |
| –18.4 | 90 (5.0) | 1,643 (9.2) | 124 (7.9) | 661 (10.0) | 2,642 (11.8) |
| 18.5–24.9 | 726 (40.5) | 9,246 (51.6) | 833 (53.3) | 3,504 (53.1) | 11,193 (49.8) |
| 25.0– | 903 (50.4) | 5,053 (28.2) | 478 (30.6) | 1,568 (23.8) | 6,484 (28.9) |
| Unknown | 73 (4.1) | 1,965 (11.0) | 127 (8.1) | 860 (13.0) | 2,143 (9.5) |
| Hospital volume | 404 (228–661) | 213 (119–368) | 266 (151–466) | 210 (119–75) | 279 (167–451) |
| –175 (tertile 1) | 284 (15.8) | 7,199 (40.2) | 482 (30.9) | 2,734 (41.4) | 6,006 (26.7) |
| 176–344 (tertile 2) | 466 (26.0) | 5,783 (32.3) | 490 (31.4) | 2,016 (30.6) | 8,026 (35.7) |
| 345– (tertile 3) | 1,042 (58.1) | 4,925 (27.5) | 590 (37.8) | 1,843 (28.0) | 8,430 (37.5) |
| Charlson comorbidity index score |  |  |  |  |  |
| 0–1 | 1,307 (72.9) | 13,570 (75.8) | 1,212 (77.6) | 5,333 (80.9) | 14,562 (64.8) |
| 2–3 | 442 (24.7) | 3889 (21.7) | 308 (19.7) | 1,120 (17.0) | 6907 (30.8) |
| 4– | 43 (2.4) | 448 (2.5) | 42 (2.7) | 140 (2.1) | 993 (4.4) |
| Comorbidity related to secondary hypertension |  |  |  |  |  |
| Diabetes mellitus | 288 (16.1) | 3,779 (21.1) | 343 (22.0) | 1,205 (18.3) | 6,750 (30.1) |
| CKD | 563 (31.4) | 1,658 (9.3) | 147 (9.4) | 529 (8.0) | 4,228 (18.8) |
| ESKD | 45 (2.5) | 527 (2.9) | 44 (2.8) | 290 (4.4) | 1,175 (5.2) |
| Sleep apnea syndrome | 65 (3.6) | 196 (1.1) | 36 (2.3) | 27 (0.4) | 461 (2.1) |
| Pheochromocytoma | 21 (1.2) | 94 (0.5) | 16 (1.0) | 23 (0.3) | 34 (0.2) |
| Primary aldosteronism | 20 (1.1) | 84 (0.5) | 16 (1.0) | 19 (0.3) | 29 (0.1) |
| Renal artery stenosis | 51 (2.8) | 187 (1.0) | 25 (1.6) | 35 (0.5) | 207 (0.9) |
| Takayasu arteritis | 1 (0.1) | 12 (0.1) | 1 (0.1) | 3 (0.1) | 32 (0.1) |
| Scleroderma | 14 (0.8) | 27 (0.2) | 3 (0.2) | 15 (0.2) | 39 (0.2) |
| Systemic lupus erythematosus | 15 (0.8) | 26 (0.1) | 4 (0.3) | 16 (0.2) | 32 (0.1) |
| Year |  |  |  |  |  |
| 2010–2011 | 320 (17.9) | 2,554(14.3) | 156 (10.0) | 1,413 (21.4) | 2,526 (11.2) |
| 2012–2013 | 376 (21.0) | 3,323(18.6) | 226 (14.5) | 1,381 (20.9) | 3,879 (17.3) |
| 2014–2015 | 394 (22.0) | 4,041 (22.6) | 287 (18.4) | 1,472 (22.3) | 5,837 (26.0) |
| 2016–2017 | 388 (21.7) | 4,297 (24.0) | 446 (28.6) | 1,301 (19.7) | 5,950 (26.5) |
| 2018–2019 | 314 (17.5) | 3,692 (20.6) | 447 (28.6) | 1,026 (15.6) | 4,270 (19.0) |

Data are numbers (percentiles) or medians (interquartile range).
BMI, body mass index; CKD, chronic kidney disease; ESKD, end-stage kidney disease.

### Trends in incidence, mortality, and urgent dialysis requirement after admission for AHT

Figure 1 illustrates the trends in the incidence of hospitalization resulting from AHT, the rates of in-hospital mortality, and urgent dialysis requirements in Japan from 2010 to 2019. As shown in Figure 1A, the absolute incidence rate remained high, ranging from 61 cases per 100,000 in 2010 to 59 cases per 100,000 admission year in 2019. Patients aged <65 years accounted for slightly less, whereas the proportion of elderly people increased. The number of female patients was greater compared with males over 10 years. The leading population of body mass was in the normal BMI group, ranged from 18.5 to 24.9, and showed an increasing trend. Notably, the incidence rates in the HHF group increased, approaching the total incidence of the remaining non-HHF group that had decreased over the recent decade.

**Figure 1.**
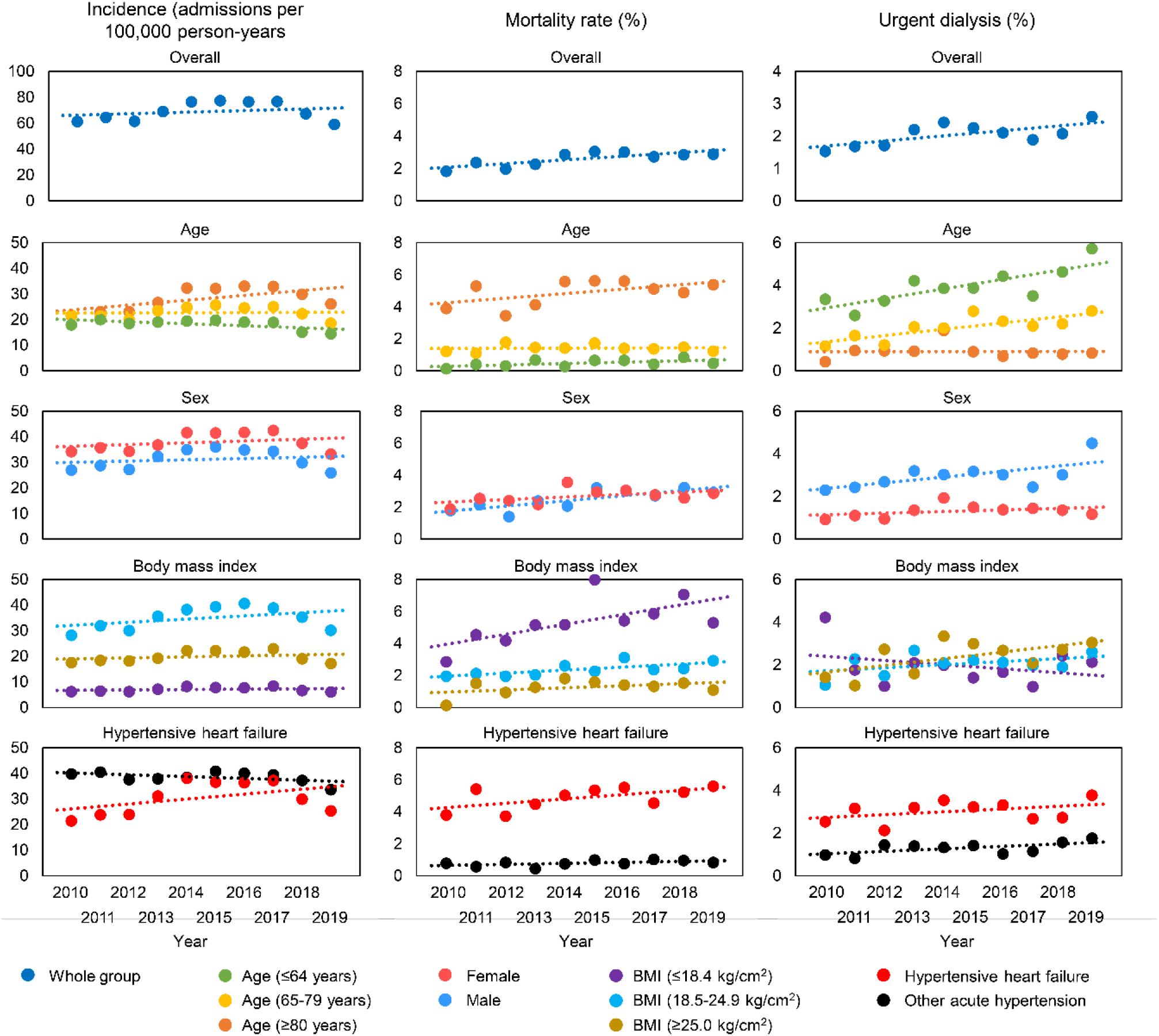
Trends in incidence, mortality, and urgent dialysis after hospitalization for acute hypertension in Japan from 2010 to 2019. Each circle represents the mean and the dashed lines represent the least squares regression lines. BMI, body mass index.

The absolute rates of all-cause death associated with AHT increased from 1.83% (95%CI, 1.40–2.40) in 2010 to 2.88% (95%CI, 2.42–3.41) (Cochran-Armitage trend test, *P* < 0.0001) in 2019 (Figure 1B). The mortality rates continued to increase for people aged <65 years, whereas that of the elderly aged >80 years markedly increased. With respect to BMI, increased mortality in the underweight group was marked and exceeded that of the normal BMI group and overweight groups over 10 years. Among the AHT spectrums, the mortality rates were much higher in the HHF-type AHT compared with the non-HHF group. The mortality rates of the HHF group increased in contrast to the gradual increase in the mortality rate of the non-HHF AHT group.

As shown in Figure 1C, the urgent dialysis rates increased from 1.52% (95%CI, 1.12–2.06) in 2010 to 2.60% (2.17–3.13) in 2019 (Cochran-Armitage trend test, *P* = 0.0071) in the subpopulation of 48,235 patients other than maintenance dialysis patients. In particular, the rates of urgent dialysis for patients under 80 years old markedly increased. The number of male patients was consistently greater than females. With respect to BMI, the rates for individuals with overweight and normal range BMI showed an overt increasing trend. The rates of urgent dialysis increased for both the HHF and non-HHF groups; however, the predominant AHT type requiring urgent dialysis was consistently HHF.

### Risk factors for in-hospital mortality in AHT

We performed a multivariable logistic regression analysis to determine risk factors for all-cause mortality following admission for AHT. As shown in Table 2, the mortality rates were higher among the elderly, males, and underweight patients. A higher number of CCI scores was associated with risk of mortality (model 1). When adjusted for causes of secondary hypertension, patients with DM, CKD, and SLE were at higher risk for death (model 2). In addition, large-volume hospitals had a lower mortality risk. In particular, the underweight group had a marked increase in risk of mortality [Odds Ratio: 1.82 (95% CI: 1.57–2.11)], whereas the overweight group tended to have reduced rates. Among the AHT spectrums, the highest mortality rates were observed for HHF, followed by MHT. No statistically significant differences were observed for the other AHT group.

**Table 2.** Factors associated with mortality in hospitalized patients with acute hypertension.

|  | Model 1 |  | Model 2 |  |
| --- | --- | --- | --- | --- |
|  | OR (95%CI) | *P* value | OR (95%CI) | *P* value |
| Age per 5 years | 1.38 (1.34–1.43) | <0.001 | 1.40 (1.35–1.44) | <0.001 |
| Female | 0.81 (0.72–0.91) | <0.001 | 0.80 (0.72–0.91) | <0.001 |
| BMI (kg/m<sup>2</sup>) |  |  |  |  |
| –18.4 | 1.76 (1.52–2.05) | <0.001 | 1.82 (1.57–2.11) | <0.001 |
| 18.5–24.9 | Reference |  | Reference |  |
| 25.0– | 0.84 (0.71–0.99) | 0.039 | 0.85 (0.72–1.01) | 0.06 |
| Hospital volume |  |  |  |  |
| –175 (tertile 1) | Reference |  |  |  |
| 176–344 (tertile 2) | 0.89 (0.78–1.02) | 0.090 | 0.88 (0.77–1.01) | 0.08 |
| 345– (tertile 3) | 0.68 (0.58–0.80) | <0.001 | 0.67 (0.57–0.78) | <0.001 |
| Year pear 1 increase | 0.98 (0.95–1.00) | 0.058 | 0.98 (0.96–1.01) | 0.2 |
| Acute hypertension spectrum |  |  |  |  |
| Malignant hypertension | 3.03 (1.12–8.23) | 0.029 | 2.75 (1.02–7.45) | 0.047 |
| Hypertensive emergency | 1.58 (0.74–3.40) | 0.2 | 1.55 (0.72–3.33) | 0.3 |
| Hypertensive urgency | Reference |  | Reference |  |
| Hypertensive encephalopathy | 1.84 (0.84–4.05) | 0.1 | 1.76 (0.80–3.86) | 0.2 |
| Hypertensive heart failure | 7.11 (3.36–15.0) | <0.001 | 6.98 (3.30–14.7) | <0.001 |
| Charlson comorbidity index score | 1.33 (1.28–1.38) | <0.001 |  |  |
| Comorbidity related to secondary hypertension |  |  |  |  |
| Diabetes mellitus |  |  | 1.03 (0.90–1.18) | 0.7 |
| CKD or ESKD |  |  | 1.97 (1.72–2.25) | <0.001 |
| Sleep apnea syndrome |  |  | 0.34 (0.13–0.93) | 0.035 |
| Pheochromocytoma |  |  | 0.79 (0.11–5.74) | 0.8 |
| Renal artery stenosis |  |  | 0.28 (0.04–2.01) | 0.2 |
| Takayasu arteritis |  |  | 1.08 (0.14–8.04) | 0.9 |
| Scleroderma |  |  | 1.89 (0.57–6.25) | 0.3 |
| Systemic lupus erythematosus |  |  | 3.88 (1.32–11.4) | 0.013 |

BMI, body mass index; CKD, chronic kidney disease; ESKD, end-stage kidney disease.

### Risk factors for urgent dialysis in AHT

After adjusting cofounders (both model 1 and 2), the rates of urgent dialysis had been still increasing. As shown in Table 3, the risk of urgent dialysis was higher among the younger, male, and those with comorbidities such as DM, CKD, and scleroderma. In addition, small-volume hospitals had a lower risk of urgent dialysis. In terms of BMI, the overweight group showed a lower risk of urgent dialysis, but not a particularly increased or decreased risk in the underweight group compared to the normal BMI group. Notably, the trend had changed dramatically over the past decade, with the overweight group having the highest rates of urgent dialysis over the other groups since 2017. The greater number of CCI scores was associated with a risk of urgent dialysis (model 1). When adjusted for causes of secondary hypertension, patients with DM, CKD, and scleroderma were at a greater risk of urgent dialysis (model 2). Among the AHT spectrums, the highest urgent dialysis rates were observed in HHF, followed by MHT.

**Table 3.**
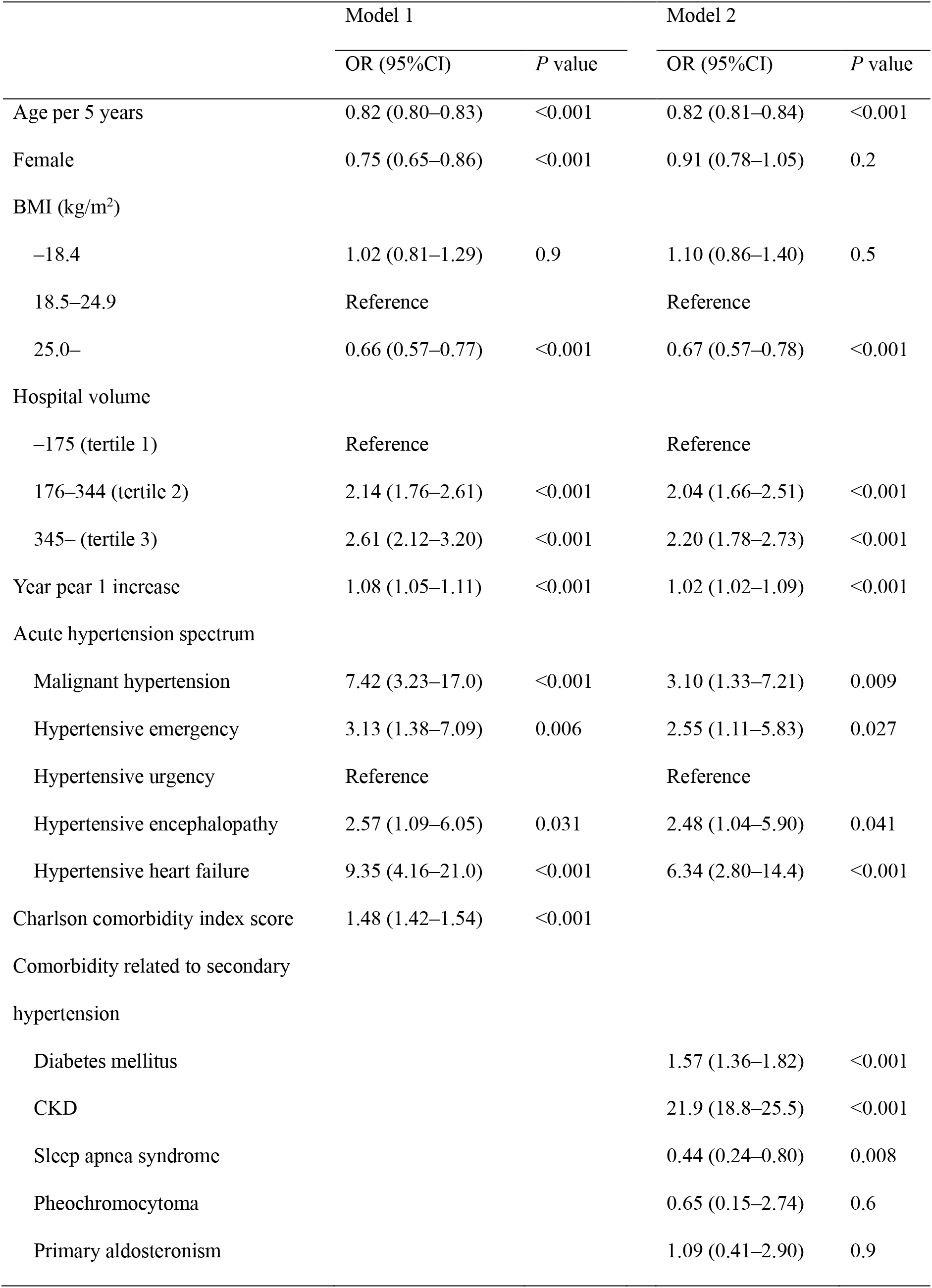
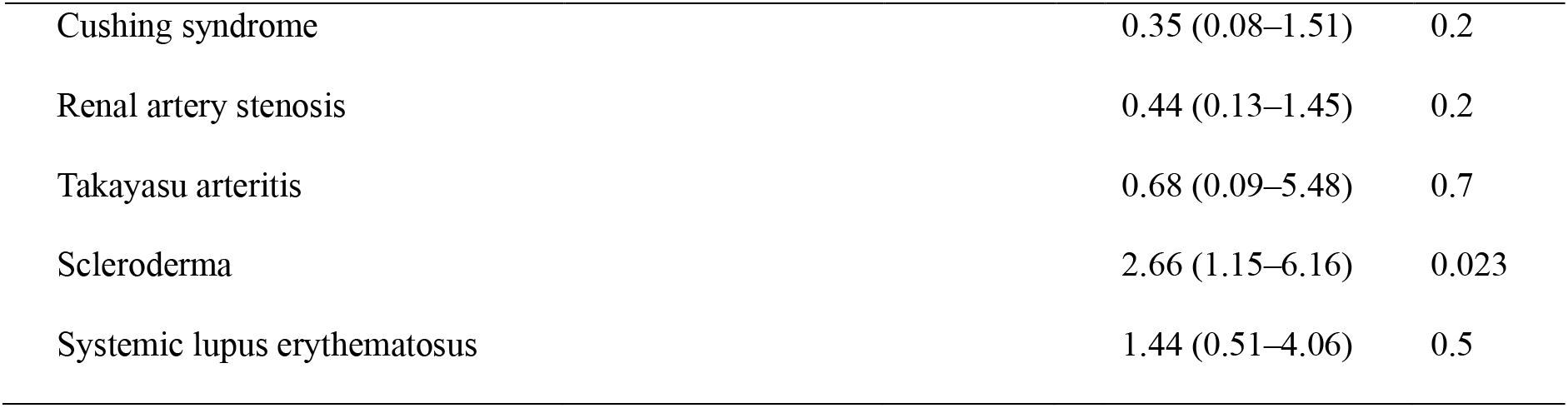
Factors associated with risk for urgent dialysis in acute hypertension and the spectrum.

### Urgent dialysis contributes to higher mortality

Next, we determined whether urgent dialysis contributes to mortality. As shown in Figure 2, there was no significant difference in the mortality risk for the ESKD group of dialysis patients compared with the control group of dialysis-independent patients. In contrast, the risk of mortality was significantly higher in the urgent dialysis group even after adjusting for confounders.

**Figure 2.**
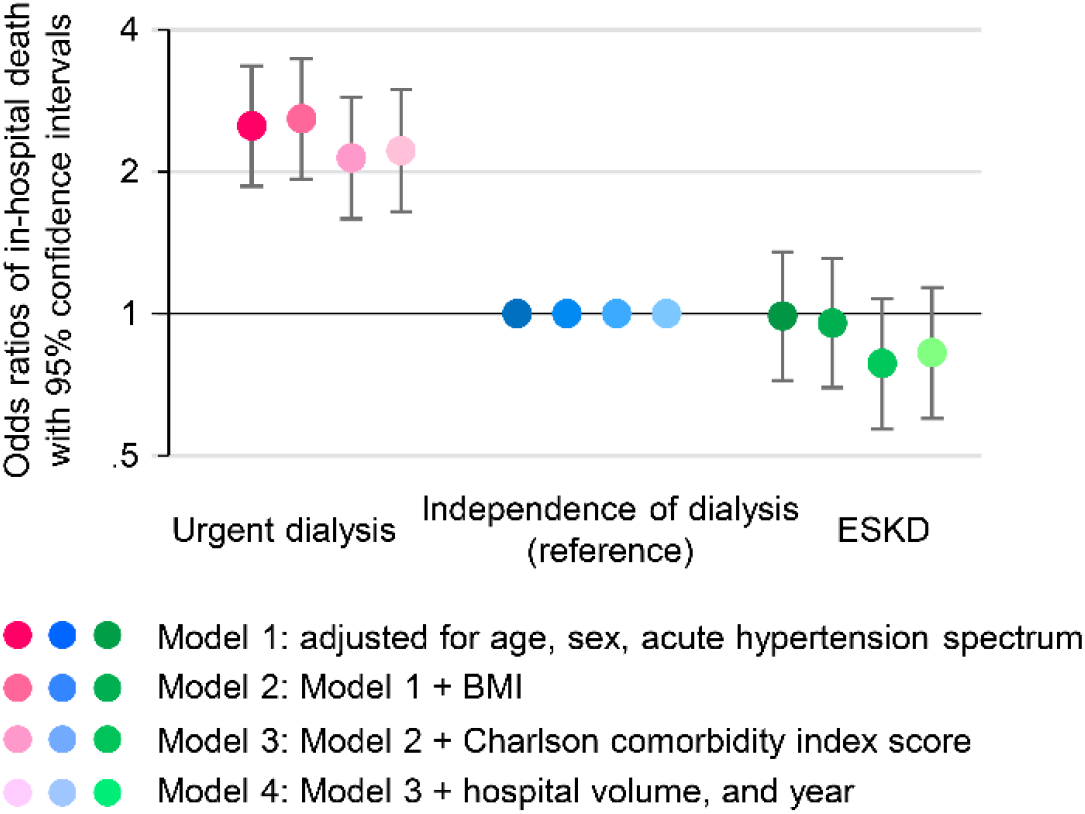
Impact of urgent dialysis on mortality after hospitalization for acute hypertension. Each circle represents the mean and the solid lines represent the corresponding 95% CI. Multivariable logistic regression models were adjusted for age, sex, acute hypertension spectrum (Model 1) and BMI (Model 2) and Charlson comorbidity index (Model 3) and hospital volume and admission year (Model 4). CI, confidence interval. ESKD, end-stage kidney disease. BMI, body mass index.

## Discussion

This population-based study clarified the nationwide increasing trends in mortality and urgent dialysis following admission for AHT from 2010 to 2019. We determined the elderly, male, lean body mass, HHF, and MHT presentations for AHT as well as the comorbidities, including DM, CKD, and SLE as risks for mortality. We also found that males, HHF-type AHT, and comorbidities, including DM, CKD, and scleroderma, were risks for urgent dialysis. The requirement for urgent dialysis was associated with greater mortality after adjusting for confounding variables. Intensive care for blood pressure and the prevention of unplanned admissions for these clinical scenarios may improve the outcomes of hypertensive patients.

This is the first report describing trends and outcomes over the most recent decade for Japanese AHT adults based on population-based nationwide datasets.^10–12^ Previous studies identified trends and outcomes of AHT in Europe and the United States;^3, 5–8^ however, these studies were based on a relatively small population with a specific area and the data from one previous decade. The more recent large-scale study updated the trends in mortality after hospitalization for AHT, but the population was limited to the elderly.^4^ The present study updated the recent national epidemiology of AHT and addressed the incidence of urgent dialysis and its impact on outcomes. We demonstrated that the crude mortality rate had been increasing since 2010, with a particular increase in the number of elderly patients, underweight patients, and HHF-type AHT. The incidence of hypertensive emergency in the United States was also reported to be increased among the elderly population.^4^ Because of an increase in patients with these traits, acute hypertension remains a “malignant” disease, despite the wider availability of anti-hypertensive drugs. In addition to DM and CKD, SLE was a strong risk factor for death in AHT. In particular, patients with SLE may develop posterior reversible encephalopathy syndrome, which has a low prevalence, but poor prognosis.^16^ Of note, we found that the mortality risk was lower in larger volume hospitals, which may be attributable to the more intensive treatment of blood pressure, congestion, and comorbidities. The reduction in mortality in larger volume hospitals is also reported after hospitalization for heart failure, acute myocardial infarction, or pneumonia.^17, 18^ Therefore, the acute-setting care of the larger hospitals may improve the outcomes of AHT patients.

We determined factors that contribute to in-hospital mortality. The risk was higher in the underweight group, whereas it was lower in the overweight group, which indicates an obesity paradox. The obesity paradox, which is a “paradoxical” decrease in mortality concomitant with increased BMI, has been shown in several populations, such as elderly, CKD, hypertension, and acute, or chronic heart failure patients.^19–21^ In patients with a progressive loss of body mass, it may be difficult to promptly find hidden congestion, if absent of an overt rise in blood pressure. Nutritional status and physical capacity profoundly affect prognosis.^22–28^ Several studies have indicated that nutrition and exercise intervention can increase muscle mass in high-risk patients with cardiovascular disease.^29–32^ Although the obesity paradox can be explained by the presence of reverse causation, such as underlying comorbidities causing weight loss, sarcopenia, and frailty, nutritional approaches to preserve metabolic reserve may improve the outcome of AHT.

The increase in HHF over the most recent decade was a substantial burden on the mortality of AHT patients. The incidence of hypertensive emergency in heart failure patients has increased in the United States.^33^ As shown in Figure 2, the requirement for urgent dialysis resulting from congestion and acute kidney injury (AKI) was linked to increased mortality after adjusting for covariates. Attempts to decrease patients requiring urgent dialysis would potentially mitigate the increasing trends in mortality. Hypertension remains one of the major reasons for developing heart failure.^34^ AKI is also an established factor contributing to cardiovascular events and death.^35^ Usual care for fluid volume and blood pressure is essential in the outpatient setting for patients with hypertension, CKD, or heart failure.

We found that the risk of urgent dialysis was greater for comorbidities, such as DM, CKD, and scleroderma, which are classically well-known risks for AHT.^36^ Scleroderma renal crisis is characterized by the abrupt onset of moderate to marked hypertension and kidney failure. Early detection and intervention are important because the outcomes remain poor, with 20-50% of patients developing end-stage renal disease.^37^ Secondary hypertension is reported to be more common in patients with hypertensive emergencies compared with the other hypertensive population.^38^ In our analysis, secondary hypertension was relatively common, especially in MHT, with SLE being a mortality risk and scleroderma associated with an urgent dialysis risk. Therefore, the risk of severe outcomes must be considered when treating patients for secondary hypertension, including autoimmune diseases.

Urgent dialysis usually requires catheter insertion or exposure to anti-coagulants. Urgent dialysis with catheter insertion potentially leads to additional complications, such as infection or bleeding, which may contribute to higher mortality.^39^ Additionally, in patients with congestion, acute correction of congestion results in all-cause and cardiovascular death.^40^ In contrast, the outcome of AHT for maintenance dialysis was favorable compared with the dialysis-independent population, despite underlying multi-morbidities. This is presumably due to good accessibility to dialysis, which is a powerful intervention for fluid overload, and the usual maintenance of body water balance usually thrice weekly in dialysis patients with HHF-type AHT. Of note, the rising trend in urgent dialysis was quite similar among the younger, male, overweight, and HHF group, indicating a specific high-risk population with these factors. CKD and progression to ESKD are associated with obesity.^41^ The effect of being overweight on years of life lost was reported to be the greatest for young adults, but decreased in the elderly.^42^ A recent large epidemiologic study of the young adult population revealed that the highest age-standardized death rates were observed for hypertension followed by obesity. Because increasing obesity-related mortality trends underlie the concerning growing global burden of metabolic diseases, intervention for this specific high-risk population may also be important.^43^

The strength of this study was the large-scale, nationally representative data for the general population of Japan.^10–12^ Previous studies have reported the validity of diagnoses and procedure records.^44^ However, our study had several limitations. First, our study lacked laboratory data, including kidney function, cardiac function, and nutritional status, in addition to imaging data. This precluded a detailed discussion related to the mechanism underlying the higher mortality in the underweight group and stratification of the mortality or urgent dialysis by kidney function. Second, the blood pressure data at admission and during hospitalization were not available. Third, the data were obtained from a single race/ethnicity. Several studies reported the ethnic variations in incidence and mortality of AHT, showing that they were higher for blacks and lower for Asians.^45, 46^ This study showed the previously undetermined trends in outcomes of AHT for Asians. Fourth, we could not analyze long-term follow-up outcomes after discharge because of the nature of the inpatient datasets. Thus, further studies are needed to clarify the long-term outcome of AHT.

### Perspective

The incidence of AHT remains high and the crude mortality has increased over the most recent decade. This is due, in part, to the increasing HHF-type scenario, aging, and the lean body mass of the AHT population. The trend of urgent dialysis has also been increasing and was associated with a greater risk of death. Usual care of latent congestion, blood pressure, and nutritional status and prevention of unplanned admissions and subsequent urgent dialysis, may potentially mitigate the premature death of hypertensive patients.

## Data Availability

The data underlying the results in this study are available from the DPC Research Institute, 4F Ginmatsu Building 1-14-11 Ginza, Chuo-ku, Tokyo 104-0061 for researchers who meet the criteria for access to confidential data.

## Acknowledgments

We would like to thank all of the study participants.

## Supplemental Materials

Figure S1. Patient flowchart

Table S1. Definition of diagnoses related to secondary hypertension based on ICD-10 codes

BMI, body mass index; CKD, chronic kidney disease; ESKD, end-stage kidney disease. Multivariable logistic regression models were adjusted for age, sex, BMI, hospital volume, admission year, acute hypertension spectrum, and Charlson comorbidity index (Model 1) or comorbidities related to secondary hypertension (Model 2).

**Novelty and Relevance**

**What Is New?**

- This study determined the nationwide trends in incidence rates, mortality, and urgent dialysis after admission for acute hypertension (AHT) from 2010 to 2019 in Japan.
- We found risk factors associated with overall death and urgent dialysis in adults with AHT and urgent dialysis was linked to a higher mortality.
- This study demonstrated clinical scenarios of AHT that contribute to greater risks of in-hospital death and urgent dialysis in hypertensive patients.

**What Is Relevant?**

- Increasing hypertensive heart failure type, aging, and lean body mass of the AHT population contribute to the rising trend in the absolute mortality rate.
- A larger hospital volume provided better survival in patients hospitalized for AHT.
- Prevention of unplanned admissions for AHT is essential for hypertensive patients.

**Clinical/Pathophysiological Implications?**

- Usual care for latent congestion, blood pressure, and nutritional status in the outpatient setting may improve the outcome of AHT.
- Prevention of unplanned admissions of adults with heart failure, chronic kidney disease, or lean body mass requiring urgent dialysis can mitigate the premature death of hypertensive patients.

